# Testing an analogue game to promote peer support and person-centredness in education for people with type 2 diabetes: a realist evaluation

**DOI:** 10.1101/2020.09.14.20185769

**Authors:** Vibeke Stenov, Pil Lindgreen, Ingrid Willaing, Henning Grubb Basballe, Lene Eide Joensen

## Abstract

**Aim:** To explore the outcomes of testing an analogue game to incorporate person-centredness and peer dialogues in group-based diabetes education targeting people with type 2 diabetes

**Design:** A quasi-experimental design inspired by realistic evaluation focusing on context, mechanisms and outcomes of the intervention

**Methods:** In March-July 2019, the game was tested among 76 people with type 2 diabetes and 17 healthcare professionals in 19 settings across nine Danish municipalities. Data consisted of audio recordings, individual and group interviews and questionnaires. Data were analysed using systematic text condensation and descriptive statistics.

**Results:** Outcomes of using the analogue game in diabetes education were: 1) a playful and relaxed atmosphere; 2) active engagement 3) reflections on diabetes-specific experiences; 4) structured and focused dialogues; 5) healthcare professionals gaining insight into the preferences and needs of participants; and 6) healthcare professionals experiencing peer dialogue as important to incorporate into education. Questionnaire responses showed that 92% of people with type 2 diabetes and 94% of healthcare professionals found that the game incorporated person-centredness and peer dialogues into diabetes education.

**Conclusion:** Implementing the analogue game as part of patient education facilitated peer support and person-centredness in a fun and playful way. Lack of time in diabetes education programmes and complicated game rules inhibited person-centeredness and peer dialogue.

**Impact:**

- The study provides novel insights into gaming as a method for promoting peer dialogue and person-centredness in diabetes education targeting people with type 2 diabetes.
- The game proved feasible as a structured tool to implement in group-based diabetes education.
- Implementing the game in diabetes education can help healthcare professionals provide diabetes education and support, which may improve quality of life and diabetes self-management skills.

## 1. INTRODUCTION

Living with type 2 diabetes (T2D) is demanding and requires lifelong self-management to prevent diabetes complications and enhance quality of life (Young-Hyman et al., 2016). Diabetes self-management education (DSME) is an essential component of care to support people with type 2 diabetes (PWT2D) in implementing self-management in their daily lives outside clinical settings (American Diabetes Association, 2020; Fan & Sidani, 2009). A global study among health care professionals (HCPs), PWT2D and their family members found that healthcare systems are poorly equipped to effectively support PWT2D (Holt et al., 2013). Although DSME was considered important, access was limited and typically not well-organized due to a lack of resources for providing psychological support (Holt et al., 2013).

Applying principles of person-centredness supports care in which PWT2D are actively involved and care that is responsive to their individual needs and preferences (Inzucchi et al., 2012; Mead & Bower, 2000). Another promising method to provide emotional support for ongoing self-management is peer support. However, effective methods and interventions to enhance peer support and person-centredness in diabetes care are needed (Joensen et al., 2016).

Use of gaming elements, such as picture cards, quotations and gamification, to motivate and engage people in non-gaming contexts encourages reflection among PWT2D, primes them to be active participants and engages them in peer dialogue during DSME (Deterding et al., 2011; Jensen et al., 2016; Torenholt, Engelund, et al., 2015; Varming et al., 2018). Furthermore, gaming elements that promote dialogue in group-based DSME improve self-management skills among PWT2D, who prefer it to traditional care (Varming et al., 2015).

### 1.1 Background

PWT2D who are more actively engaged in their care report better clinical outcomes, higher quality of life, healthier behaviours and enhanced self-management skills (Hibbard et al., 2007). Despite attempts to define person-centredness (Mead & Bower, 2000; Pulvirenti et al., 2014), no agreed-upon definition has yet been accepted, resulting in diverse uses of the term (McCance et al., 2011). However, the European Association for the Study of Diabetes and the American Diabetes Association have defined a person-centred approach as “providing care that is respectful of and responsive to individual patient preferences, needs and values” (Inzucchi et al., 2012, p. 1364). Peer support is a method for providing diabetes-specific social support and creating person-centeredness (Boothroyd & Fisher, 2010; E. B. Fisher et al., 2015), and it has been found to be an effective method to provide support for ongoing self-management in PWT2D (Funnell, 2010; Heisler, 2010; van Dam et al., 2005).

Despite evidence of the positive effects of person-centredness and peer support and the intention of HCPs to include these approaches, implementing them in practice is challenging (Odgers-Jewell et al., 2015; Stenov et al., 2017). A major barrier is the shift that HCPs must make from being didactic experts with limited time for peer interaction to being a facilitator applying a collaborative approach (L. Fisher et al., 2017). In particular, many HCPs find it challenging to adopt participatory methods in DSME because they lack required experience and training (Holt et al., 2013; Stuckey et al., 2015).

Game design approaches support active involvement and face-to-face peer interactions in healthcare settings (Gauthier et al., 2019). Studies have demonstrated promising outcomes of playing educational games on a variety of factors in PWT2D, such as motivation for behaviour change and diabetes outcomes (Deen & Schouten, 2011; Gauthier et al., 2019; Shaffer, 2007). However, fully integrated and structured games are currently designed primarily for digital use and as educational media to provide information or enhance self-management skills related to, for example, the relationship between food, insulin, physical exercise and blood glucose levels (Gauthier et al., 2019; Lazem et al., 2016). Despite rapid growth in the number of initiatives employing digital and educational games in diabetes care, many games are designed for children or adolescents with diabetes (Bochennek et al., 2007; de Vette et al., 2015). Although games may have the potential to improve diabetes self-management (Bochennek et al., 2007), to the best of our knowledge, no previous study has investigated whether and how an analogue game can facilitate person-centredness and peer dialogue in DSME.

## 2. THE STUDY

### 2.1 Aim

The aim of this realist evaluation study was to explore the outcomes of using an analogue game aimed at incorporating peer support and patient-centredness in group-based DSME targeting PWT2D.

### 2.2 Study Design

A quasi-experimental design inspired by the realist evaluation approach was used (Ray & Nick, 2014). This approach was selected because it explores concrete demonstrations of hypothesized contexts, mechanisms and outcomes (CMOs) of an intervention implemented in a specific setting (Ray & Nick, 2014). The realist evaluation approach applies a programme theory to understand how, for whom and under what conditions a specific intervention will work and which outcomes it will produce (Ray & Nick, 2014).

### 2.3 The analogue game

#### 2.3.1 Game design

The development of the analogue game was inspired by design thinking (Brown & Wyatt, 2010), which comprises three phases: ideation, development and implementation (Dolmans & Tigelaar, 2012). Game design reflected the first two phases, whereas the realist evaluation of the game reflects the implementation phase. The game was designed in January 2018-January 2019 in a partnership between designers from Copenhagen Game Lab, who specialize in designing and conducting iterative co-creative game design processes, and researchers from Steno Diabetes Center Copenhagen with professional backgrounds in user-driven innovation and psychology, communication, public health science and nursing. In addition, 37 PWT2D, four HCPs, a diabetes psychologist and a graphic designer were involved in the development phase, which included multiple workshops including 3-12 PWT2D and using various methods to promote ideation and prototype development (Appendix 1).

#### 2.3.2 Expected mechanisms and outcomes

The primary aims of the analogue game were to: 1) create a psychologically safe environment allowing PWT2D to systematically engage in peer dialogues about life with T2D and 2) provide HCPs with insights into the challenges, needs and preferences of PWT2D participating in the DSME programme.

The game was informed by the key concepts of person-centredness (Mead & Bower, 2000) and peer support (E. B. Fisher et al., 2015), which are underpinned by the theories of empowerment (Anderson & Funnell, 2005) and social learning (Bandura, 1977). The program theory identifies the expected mechanisms and outcomes of the analogue game, as well as contextual conditions that influenced the expected mechanisms and outcome (Table 1). The program theory also guided data collection and analysis.

**Table 1.** Programme theory for the game intervention

| Context | Mechanisms | Outcomes |
| --- | --- | --- |
| <u>Educational environment</u><br>The game:<br>Game mechanisms need to match the learning preferences of the group<br>The game needs to be played at the “right” time (neither too early nor too late in the DSME programme)<br>Allowing enough time in the DSME programme for the game to be played<br>HCPs:<br>HCPs need to be supportive of an open-minded dialogue and confident in the context of active engagement of PWT2D | <u>Gamification</u><br>Designing game elements that create:<br>Playful, competitive and rewarding activities, including the possibility of winning or losing the game<br>A simulation of various applicable coping strategies for life with T2D<br><u>Structure</u><br>Creating a self-facilitating game flow that:<br>Alternates between game elements and reflection phases<br>Engenders discussions and invites players to prioritize good discussions over game play<br><u>Fictitious personas</u><br>Designing fictional, relatable personas that enable:<br>Normalization and non-judgmental mirroring<br>Emotional distancing (by linking diabetes challenges to personas rather than oneself or other players)<br><u>Theme cards</u><br>Combining a title, a picture and a quote that:<br>Inspire and prompt reflections upon life with T2D<br>Give PWT2D multiple choices to address various needs and share relevant experiences | <u>For PWT2D</u><br>Experiencing a playful and relaxed atmosphere leading to open-mindedness in the DSME programme<br>Being actively engaged in the DSME programme<br>Reflecting and discussing diabetes-specific experiences<br>Participating in structured and focused dialogue<br><u>For HCPs</u><br>Gaining insight into the daily lives, attitudes, wishes, needs, challenges and preferences of PWT2D<br>Creating a legitimate framework for peer dialogue and person-centredness, strengthening the application of both in DSME programmes |

#### 2.3.3 Game content

The final version of the game consists of visual and tangible materials, such as laminated cards with illustrations and quotes, as well as game elements intended to stimulate reflection and dialogue among PWT2D and engage them in a fun and playful way. The game is played by a group of three to five PWT2D, each of whom plays as a fictitious persona with T2D. Each player selects diabetes-related theme cards that best fit their game persona; cards include tips and advice in the four domains of diet, exercise, medication and social relations with family and friends. The better the theme card fits the persona, the more points the player receives. The players then discuss their own experiences, challenges and needs in life with T2D, based on the selected cards. Table 2 provides an overview of the game content.

**Table 2.**
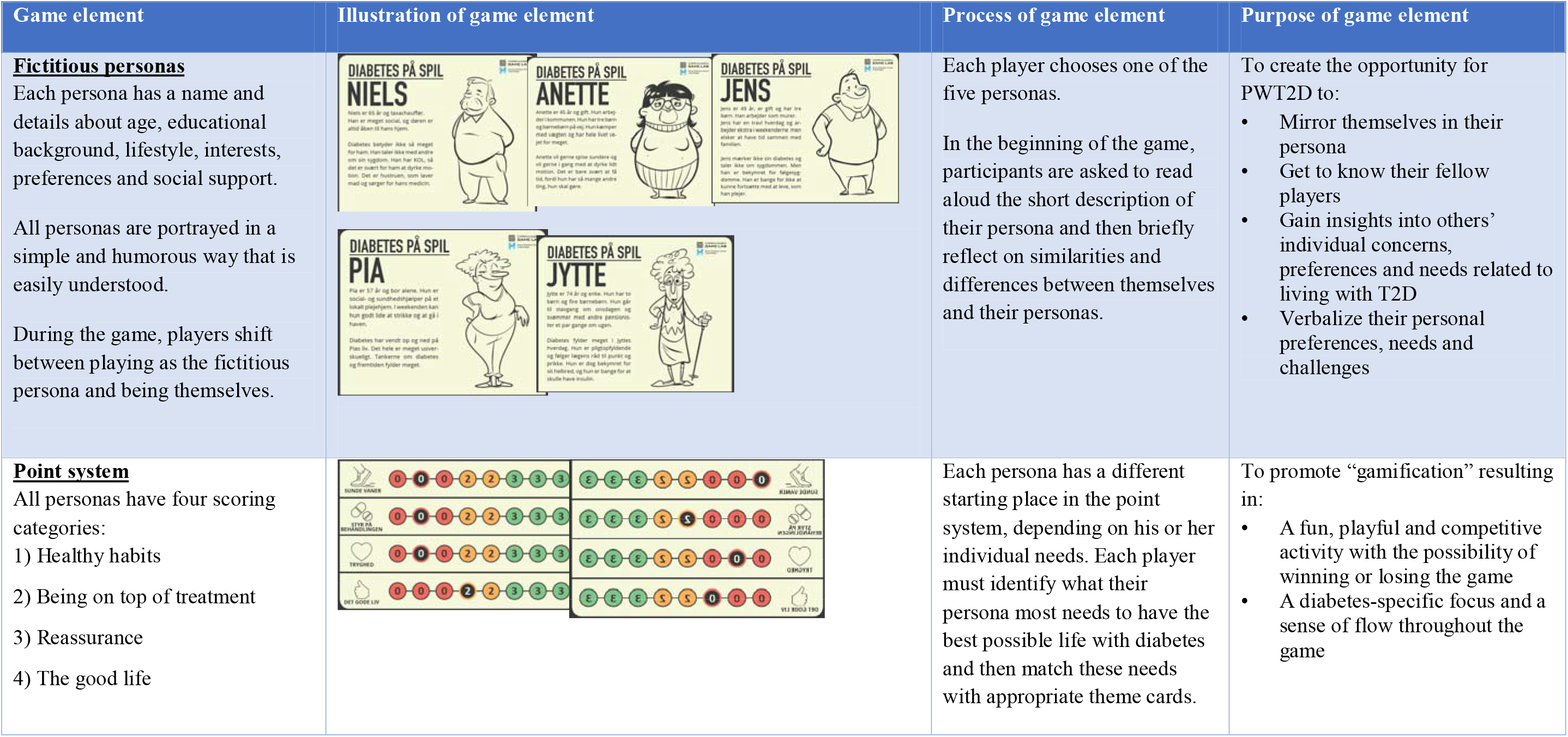
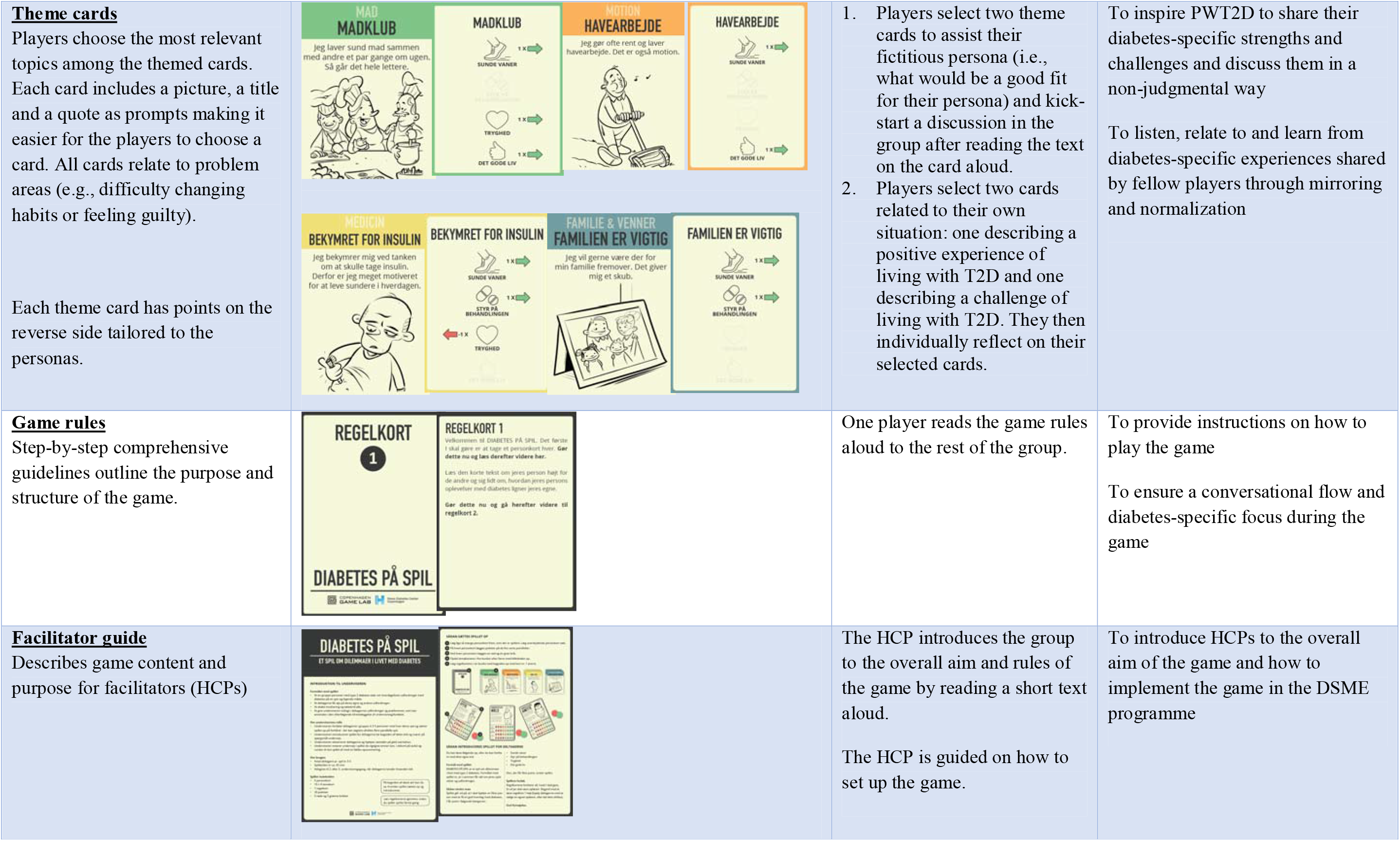
Overview of game content

| Game element | Illustration of game element | Process of game element | Purpose of game element |
| --- | --- | --- | --- |
| **<u>Fictitious personas</u>**<br>Each persona has a name and details about age, educational background, lifestyle, interests, preferences and social support.<br>All personas are portrayed in a simple and humorous way that is easily understood.<br>During the game, players shift between playing as the fictitious persona and being themselves. |  | Each player chooses one of the five personas.<br>In the beginning of the game, participants are asked to read aloud the short description of their persona and then briefly reflect on similarities and differences between themselves and their personas. | To create the opportunity for PWT2D to:<br>• Mirror themselves in their persona<br>• Get to know their fellow players<br>• Gain insights into others' individual concerns, preferences and needs related to living with T2D<br>• Verbalize their personal preferences, needs and challenges |
| **<u>Point system</u>**<br>All personas have four scoring categories:<br>1) Healthy habits<br>2) Being on top of treatment<br>3) Reassurance<br>4) The good life |  | Each persona has a different starting place in the point system, depending on his or her individual needs. Each player must identify what their persona most needs to have the best possible life with diabetes and then match these needs with appropriate theme cards. | To promote “gamification” resulting in:<br>• A fun, playful and competitive activity with the possibility of winning or losing the game<br>• A diabetes-specific focus and a sense of flow throughout the game |
| **Theme cards**<br>Players choose the most relevant topics among the themed cards. Each card includes a picture, a title and a quote as prompts making it easier for the players to choose a card. All cards relate to problem areas (e.g., difficulty changing habits or feeling guilty).<br>Each theme card has points on the reverse side tailored to the personas. |  | 1. Players select two theme cards to assist their fictitious persona (i.e., what would be a good fit for their persona) and kick-start a discussion in the group after reading the text on the card aloud.<br>2. Players select two cards related to their own situation: one describing a positive experience of living with T2D and one describing a challenge of living with T2D. They then individually reflect on their selected cards. | To inspire PWT2D to share their diabetes-specific strengths and challenges and discuss them in a non-judgmental way<br>To listen, relate to and learn from diabetes-specific experiences shared by fellow players through mirroring and normalization |
| **Game rules**<br>Step-by-step comprehensive guidelines outline the purpose and structure of the game. |  | One player reads the game rules aloud to the rest of the group. | To provide instructions on how to play the game<br>To ensure a conversational flow and diabetes-specific focus during the game |
| **Facilitator guide**<br>Describes game content and purpose for facilitators (HCPs) |  | The HCP introduces the group to the overall aim and rules of the game by reading a short text aloud.<br>The HCP is guided on how to set up the game. | To introduce HCPs to the overall aim of the game and how to implement the game in the DSME programme |

### 2.4 Participants and setting

During five months in 2019, the game was tested in 19 settings in nine municipalities across Denmark comprising urban and rural settings in different geographical locations. Inclusion criteria for PWT2D were: a diagnosis of T2D, age ≥ 18 years, and no comorbid psychosis or dementia. Seventy-seven PWT2D and 17 HCPs participated by playing (PWT2D) or facilitating (HCPs) the game, completing questionnaires and being interviewed. PWT2D were interviewed in focus groups and HCPs were interviewed in pairs or individually.

### 2.5 Data collection

#### 2.5.1 Game tests and observations

Tests lasted 1-1½ hours each and were audio recorded. Field notes were created based on observations of the game tests, using a semi-structured observation guide focusing on body language, atmosphere, dialogue among peers, physical surroundings and the physical layout of the game (Spradley, 2016). Audio recordings were transcribed verbatim and primarily used to identify whether and how hypothesized CMOs were reflected in practice, including how and to what extent PWT2D shared their experiences with peers, expressed their needs and challenges and were actively involved in the game.

#### 2.5.2 Interviews with PWT2D and HCPs

Focus group interviews with PWT2D and individual or dyadic interviews with HCPs explored their experiences and appraisal of game play and outcomes (Koch & Vallgårda, 2008) and investigated the game’s usability, applicability and implementation potential. A semi-structured interview guide related to specific CMO game configurations included three topics: 1) the experience of playing the game and perspectives on its usability; 2) the potential for discussing meaningful diabetes-specific topics with co-players during the game; 3) the potential for peer dialogues among PWT2D playing the game. Interviews were audio recorded and transcribed verbatim.

#### 2.5.3 Questionnaires

Game evaluation questionnaires were developed separately for PWT2D and HCPs, guided by the programme theory of the game. The PWT2D questionnaire included four topics: 1) sociodemographic data; 2) overall experience of the game and its elements; 3) experience of peer dialogues occurring during the game; 4) experienced outcomes of playing the game. It was pilot tested with 13 PWT2D at Steno Diabetes Center Copenhagen to ensure that questions were legible, relevant and easily understood and to assess the time required for completion. The questionnaire for HCPs mirrored the topics of the questionnaire for PWT2D and also included items about the implementation potential and structure of the game. To minimize information bias and enhance the response rate, participants were individually asked to complete questionnaires immediately after game tests and before interviews.

### 2.6 Data analyses

Questionnaire data were analysed in SPSS Statistics 25 (Marija, 2011) using descriptive statistics. Furthermore, to explore differences in game experiences and satisfaction related to gender, educational level, cohabitation status and specific settings and participants, post hoc analysis with Chi-square tests was performed, with statistical significance set at p < 0.05.

To identify game mechanisms, field notes and transcripts from the game tests and interviews were analysed using systematic text condensation (Malterud, 2012), which consists of the following steps: 1) reading through the material to identify preliminary themes; 2) identifying and developing meaning units; 3) systematically abstracting meaning units; 4) reconceptualising the data and generating concepts and descriptions (Malterud, 2012). Qualitative data were organised and analysed in NVivo 12 Pro (Troy, 2018).

### 2.7 Validity and rigour

Study rigour was ensured by the development of a program theory guiding data collection and analysis, with the aim of systematically exploring mechanisms, contextual conditions and self-reported participant outcomes related to the game. Furthermore, study validity and rigour were pursued by the use of multiple data collection methods to explore different aspects of usability of the game as a whole and of its elements (Patton, 2002). Including nine municipalities in varied geographical locations across Denmark ensured that participants of various backgrounds were represented. Finally, data coding was conducted by two researchers and a research assistant, and all authors participated in interpreting and discussing subthemes and themes until agreement on the final themes and comprehensive understanding was reached. The varying professional backgrounds of the authors in public health, psychology and nursing enriched discussions, provided diverse data interpretation perspectives and were viewed by the authors as enhancing trustworthiness.

### 2.8 Ethical considerations

The study was conducted in accordance with the Helsinki Declaration and approved by the Danish Data Protection Agency (VD-2018-157). No ethical approval from the Danish Health Research Ethics Committee was required (http://en.nvk.dk/how-to-notify/what-to-notify). Participants received verbal and written information on the study before giving written informed consent to take part in game tests. All collected data were anonymized and handled confidentially in accordance with Danish legislation.

## 3. FINDINGS

Table 3 displays the characteristics of PWT2D and HCPs. Six outcomes of using the analogue game in DSME were identified and exemplified the expected outcomes of the programme theory (Table 1). Table 4 includes qualitative data illustrating mechanisms that enabled or inhibited achieving desired outcomes and contextual conditions that affected outcomes and mechanisms. The six outcomes are described through qualitative and quantitative data. Data from PWT2D and HCPs questionnaires are presented in Table 5.

**Table 3.** Participant characteristics

|  | **PWT2D**<br>**N=76** | **HCPs**<br>**N=17** |
| --- | --- | --- |
| Gender, female % (n) | 47 (37) | 100 (17) |
| Age in years, mean (range) | 64 (42-89) | 45 (25-65) |
| T2D duration in years, mean (range) | 7 (0.1-60.0) |  |
| No comorbidities, % (n) | 56 (40) |  |
| ≥ 1 comorbidity, % (n) | 44 (31) |  |
| Chronic obstructive pulmonary disease | 19 (6) |  |
| Depression | 16 (5) |  |
| Osteoarthritis | 42 (13) |  |
| T2D treatment, % (n) |  |  |
| Oral medication | 85 (60) |  |
| Insulin | 18 (10) |  |
| Education, % (n) |  |  |
| Primary or lower secondary | 24 (16) |  |
| Upper secondary | 1 (1) |  |
| Vocational training | 36 (24) |  |
| Short post-secondary education (< 3 years) | 11 (7) |  |
| Post-secondary education (≥ 3 years) | 20 (13) |  |
| University post-secondary education (≥ 5 years) | 8 (5) |  |
| Experience working with PWT2D in years, mean (range) |  | 7 (0-30) |
| Profession, n (%) |  |  |
| Dietitian |  | 19 (3) |
| Physiotherapist |  | 19 (3) |
| Nurse |  | 62 (11) |

**Table 4.** Mechanisms and contextual conditions affecting outcomes

| Outcomes |  |  |
| --- | --- | --- |
| For PWT2D | Enablers | Inhibitors |
| A playful and relaxed atmosphere | <u>Playful, competitive and rewarding activities enhanced a trustful atmosphere (mechanism):</u><br>P2: Playing a game creates something different when you speak together (...) It simply adds some playfulness and cheerfulness and it doesn't harm at all.<br>P3: I think it opens for more vulnerable issues, which can be difficult to talk about and this gets you closer together and makes you safer within the group (focus group 9)<br>HCP: "Now we must praise the winner and give a round of applause"<br>P6: "Did I win?"<br>P5: "You had one more point"<br>P6: "Should I stand up?" [Laughter within the group] (test 4)<br><u>Playing at the "right" time in the DSME programme facilitated cohesion (contextual condition):</u><br>P3: "It would probably have been different if we had to do it [play the game] in the beginning."<br>P4: "Then, I think, it wouldn't have worked at all."<br>P3: "It would definitely have been different."<br>P4: "Then you would've had to be extremely open."<br>P3: "It's important that you know each other a bit." (focus group 3) | <u>Judgmental comments inhibited a relaxed and open dialogue (contextual condition):</u><br>P1: "I have no problem with that [exercising outside]. Unless it's raining." A co-player responded in a negative tone: "You could simply wear a rain suit." (test 13). |
| Active engagement | <u>Game structure framed conversational flow among PWT2D (mechanism):</u><br>"I can see some potential in this game, as it initiates some dialogue. So, everyone says something. I think that's one of its strengths." (HCP, municipality D).<br>"It is very much the ones who usually do not say much that open up more [when playing the game]. He [one of the participants] is a very quiet person, but he is definitely saying more during the game (...). I observed that she [another participant] was a bit grumpy in the beginning and then during the game she relaxed a bit and became engaged." (HCP, municipality B). | <u>Complicated game content and structure inhibited active engagement (mechanism and contextual condition):</u><br>"It's problematic with a lot of text for participants with dyslexia. The text in the game is very heavy, and I think that's a disadvantage." (HCP, municipality I)<br>"I think it's too difficult for this group of participants. They simply cannot reflect on a high enough level." (HCP, municipality H).<br>"I think it's a problem. It was hard for her [a member of an ethnic minority unable to speak Danish fluently] to translate it and it was difficult for her to explain herself." (HCP, municipality A). |
| Reflections on diabetes-specific experiences | <u>Emotional distancing by linking diabetes challenges to personas enabled articulation of reflections (mechanism):</u><br>P1: "He [the persona] probably doesn't look much like me. Diabetes is relatively new to me, but, of course, at home, we discuss it. It's difficult for him [the persona] to exercise. It's probably easier for me, but it's easy to find excuses for not doing it, too. My family [son and girlfriend] can eat exactly what they want. They don't gain weight at all, but I do. Ten years ago, I was also very slim, but suddenly, it changed. I work every day, but I don't get my pulse up. It's difficult finding the time. I work 12 hours a day." (test 10).<br>P1: "You get a fictitious persona that you begin with and then afterwards put yourself in it. I think that very good. It's easy to talk about a fictional person, but what I really like about this is that you become more aware yourself and can share your own experiences too and then hear what others are telling. At our table, we were very open about everything. We asked each other a lot of questions."<br>P2: "But also, sharing some ideas to what you can do."<br>P3: "It's also the first time that we are in groups and it contributes automatically to more talking."<br>P4: "I think I got the sense that when you initially play the fictitious persona then you put other things at the table." (focus group 3). | <u>Confusing game rules led to uncertainty about how to play (mechanism):</u><br>P1: The challenge is matching the theme cards with the fictitious persona, and there is a challenge in figuring out what fits for Charlotte, Pia and Niels [names of the personas] [...] It's difficult, because there isn't really any information about the persona and then, you don't really know what to choose and you have to imagine [what they need]. The other [challenging] thing is knowing, when to talk about yourself in all of this. I think it was easier just talking about yourself." (focus group 1). |
| Structured and focused dialogues | <u>Playing the fictitious persona structured an open-minded dialogue (mechanism):</u><br>“[reading the text from the persona aloud] It doesn’t look like me, the first part [...] but the rest is spot on. I’ve struggled with my weight my entire life. My persona wants to eat more healthily and exercise, but it’s difficult finding the time [...] That fits me very, very well. I think it’s difficult to find time, but I have prioritised it [exercising] now, but I’m about to break down so I must find the balance. I have to say, diabetes means a lot, too [...] I’m afraid that it [diabetes management] suddenly doesn’t matter [to me]. I can feel it sneaking up on me. I have so many years left to constantly take care of my diabetes, and if I fall into a trap now, after not even a year, I’m terrified and afraid of losing control.”(setting 18).<br>P1: “I think it was a really good game. We have different lives and different approaches, but anyway you can recognise a lot of what has been said by others. It’s nice to hear that you not are the only one who has these thoughts (...) struggling with different things. Even though it’s not exactly the same stuff you are struggling with, it’s all about the disease, anyway. It’s nice that this game opens for sharing issues on the level you choose.” (focus group 2).<br><u>Choosing between different topics focused on diabetes supported sharing experiences (mechanism):</u><br>“It outlines a framework, right? This is where we are, and this is what we’re discussing. Actually, I think that they were good at being within the framework and giving each other space, while at the same time expressing how they felt themselves. So, the game absolutely facilitates them being more engaged.” (HCP, municipality I)<br>“The themed card about TV without candy. It’s because I’m very good at eating healthy through the day and sometimes I forget to eat, and when it’s evening then there this thing about cravings. It can be crazy. It’s good that I don’t have anything in the house then I would have it all. I think that’s tough.” (test 5) | <u>Lack of time rushed the peer-dialogue (contextual condition):</u><br>“I also felt pressured by the time factor, and the dialogue was, in fact, important. It’s not a waste of time [playing the game] so to speak, but how much do you have to push [the game forward]? And how important is it to play the entire game? Yes, that [figuring out] was a challenge.” (HCP, municipality I) |
| For HCPs: | <u>Enablers:</u> | <u>Inhibitors:</u> |
| Healthcare professionals gained insights into people with type 2 diabetes preferences and needs | <u>Limited educator talk increased time for peer dialogue (contextual condition):</u><br>“I think it was really nice to walk around and observe them because it gave me some knowledge about what their major challenges really are.” (HCP, municipality D)<br>“The fictitious persona works really well in promoting insight into each participant’s challenges and preferences, even though they don’t find themselves similar to the persona, they just explain how they were unlike.” (fieldnotes, municipality A)”.<br>“The issue about guilt and shame. Two of the women were pretty touched by it, right. I knew that one of them was vulnerable, but not that severely. Much of the stuff came up by playing the game. And when it comes up like that, then it means a lot to them. It’s good to know- you get to know them in a different way.” (HCP, municipality I) | <u>HCPs’ uncertainty about how to facilitate dialogue inhibited dialogues (contextual condition):</u><br>P 6: “Well, it [blood sugar] is stable when measuring it the next day.”<br>HCP2: “Yes. So you see the connection to whole grain and greens.”<br>P 6: “Yes, but it [blood sugar] is still elevated at the end of the day.”<br>HCP2: “I see. And how much?”<br>P 6: “Well, it goes beyond 10.”<br>HCP2: “And what does that make you think?”<br>P 6: “I don’t understand why it happens.”<br>HCP2: “And how late in the day does it happen?”<br>P 6: “After I have my dinner, my greens. Breakfast is fine.”<br>HCP2: “And how long after dinner is it?”<br>P 6: “An hour and a half.”<br>HCP2: “So, that’s when your blood sugar is the highest?”<br>P 6: “Yes” (test 16). |
| Healthcare professionals experienced peer dialogue as important to incorporate in education | <u>Playing the game created solidarity and collectiveness (contextual condition):</u><br>“[...] It [the game] creates a kind of collectiveness. And building up relationships takes time and the game supports that.” (HCP, municipality D)<br>“I could feel that it [the game] brought them closer together. I could already feel that as the game went along. That it created a kind of solidarity. It was an eye opener how much they actually needed to tell and talk together with equally minded people.” (HCP, municipality I) | <u>Used too much time in understanding the game (contextual condition):</u><br>“If we have to spend one whole session on this [playing the game], and this might be a presumption of mine, then I’m a bit nervous that some [PWT2D] will be disappointed [...] they want facts [...] and [they] prefer the blackboard and getting some handouts.” (HCP, municipality A)<br>“If I compare how much time, if we really are supposed to have to the dialogue and reflections by playing the game, then we need double time, and then I think we could have the dialogue in another way in much shorter time.” (HCP, municipality H) |

**Table 5.** PWT2D and HCP experiences of game play

|  | % (N) |
| --- | --- |
| **People with T2D** |  |
| Overall perception of the analogue game, good or very good | 92 (70) |
| The game was fun to play, agree or strongly agree | 81 (60) |
| I experienced a good atmosphere during the game, agree or strongly agree | 99 (74) |
| Game topics were relevant to everyday life with T2D, agree or strongly agree | 86 (65) |
| The fictitious persona seemed authentic, agree or strongly agree | 86 (62) |
| Playing the fictitious persona made me think of own life with T2D, agree or strongly agree | 76 (58) |
| The point system in the game was easy to understand, agree or strongly agree | 76 (57) |
| The game rules were easy to understand, agree or strongly agree | 84 (63) |
| I experienced a sense of collectiveness with others who have diabetes, agree or strongly agree | 83 (63) |
| We talked well together during the game, agree or strongly agree | 96 (71) |
| We listened to each other during the game, agree or strongly agree | 96 (72) |
| I heard about others' experience with diabetes during the game, agree or strongly agree | 83 (62) |
| I talked with others about diabetes during the game, agree or strongly agree | 88 (67) |
| **HCPs** |  |
| Overall perception of the analogue game, good or very good | 94 (16) |
| Playing the game ensured that everyone got a chance to talk, agree or strongly agree | 100 (16) |
| The game encouraged the PWT2D to talk more than usual in the DSME, agree or strongly agree | 59 (10) |
| The game was fun to play, agree or strongly agree | 81 (13) |
| I experienced a good atmosphere during the game, agree or strongly agree | 100 (16) |
| I heard about participant experiences during the game, agree or strongly agree | 88 (15) |
| Game topics were relevant and in keeping with the content of DSME, agree or strongly agree | 94 (16) |
| The fictitious persona seemed trustworthy, agree or strongly agree | 81 (13) |
| The point system in the game was easy to understand, agree or strongly agree | 63 (10) |
| The game rules were easy to understand, agree or strongly agree | 81 (13) |
| The game encouraged dialogue within the group, agree or strongly agree | 76 (13) |
| I experienced that the PWT2D listened attentively to each other during the game, agree or strongly agree | 94 (16) |
| Playing game resulted in too little time for other activities in the programme, agree or strongly agree | 62 (10) |
| It was difficult to end the conversations during the game, agree or strongly agree | 50 (8) |
| The game promoted useful knowledge on the experience and view on diabetes of each participant with T2D, $\geq 5$ on a VAS scale of 0-10 | 88 (15) |
| I will use the analogue game in my future work, $\geq 5$ on a VAS scale of 0-10 | 88 (15) |
| I will recommend the game to a colleague, $\geq 5$ on a VAS scale of 0-10 | 88 (14) |

### 3.1 A playful and relaxed atmosphere

#### 3.1.1 Playful, competitive and rewarding activities enhanced an atmosphere of trust

The point system of the game was perceived as a fun and entertaining activity that promoted laughter and a competitive spirit among players, which may have paved the way to a relaxed atmosphere of trust. Participants demonstrated a willingness to share their T2D experiences that became easier to talk about, as described by one HCP:

> Everyone says something and there was a really good atmosphere. They joke with each other. This is what the game can promote. It can lighten up the mood a bit. It’s still a game, and a game must be fun. You must try winning the game, a kind of a gaming spirit. Then you forget how serious everything is, right? (HCP, municipality B)

In the survey, 81% (60) of PWT2D and 81% (13) of HCPs agreed or strongly agreed that the game was fun to play. In addition, 99% (74) of PWT2D and 100% (17) of HCPs reported that they experienced a good atmosphere during the game (Table 5).

#### 3.1.2 Playing at the “right” time in the DSME programme facilitated cohesion

It was crucial to play the game at an appropriate time to create and maintain the level of trust and openness PWT2D needed to share experiences as part of the game: “Playing the game in the second or third session [of the DSME programme] is best, because then they experience a sense of emotional cohesion, which gives [them] a chance to talk about deeper topics before the programme ends.” (HCP, municipality A)

#### 3.1.3 Judgmental comments inhibited a relaxed and open dialogue

During game play, participants occasionally made judgmental or negative comments to each other, leading to unproductive group interactions. Blaming and shaming between players may have hindered an open atmosphere of trust:

> P2: […] I think about the consequences of my illness often, but I trust my doctor. And occasionally, I take a day off from my diabetes [title of a theme card]. It’s liberating.
>
> P3: That’s bloody silly. I hope you get minus points for that.” (game test, municipality G)

### 3.2 Active engagement

#### 3.2.1 Game structure framed conversational flow among PWT2D

The game functioned as an icebreaker and motivated participants to engage actively. The HCPs pointed out the high level of participation among players as a key benefit of the game, encouraging more reticent PWT2D to speak up: “It’s very much the ones who usually don’t say much who open up more. He [one participant] is a very quiet person, but he undoubtedly speaks up more during the game.” (HCP, municipality B)

According to some HCPs, the involvement of all participants occurred naturally due to the game structure, as opposed to standard DSME in which including everyone in the dialogue largely depends on HCP’s facilitation skills. One HCP explained how the game rules and instructions guided the conversational flow among all group participants:

> “We’re trained in dialogue-based education, so usually, we steer the conversation a bit, but in the game, you don’t have to […] because none of the participants dominated [the game]. You’re used to guiding a couple of group participants to make room for each other. The game forced the participants to make sure everyone got a say.” (HCP, municipality E)

Questionnaire responses showed that 100% (17) of HCPs agreed or strongly agreed that playing the game increased the likelihood of all participants having a chance to talk. Moreover, 96% (72) of PWT2D and 94% (16) of HCPs agreed or strongly agreed that PWT2D listened to each other during the game (Table 5).

#### 3.2.2 Complicated game content and structure inhibited active engagement

A few HCPs found that the game was unappealing to some PWT2D due to different learning preferences: “Some participants find it more difficult than others to understand the game rules and the various quotes on the cards. Not everyone has the same immediate understanding of the game” (HCP, municipality A). In a similar vein, other HCPs noted that the amount of text that must be read aloud to play the game was too difficult for some people due to dyslexia or inability to read Danish fluently.

Eighty-four percent (63) and 81% (13) of surveyed PWT2D and HCPs respectively agreed or strongly agreed that the game rules were easy to understand, while 76% (57) of PWT2D and 63% (10) of HCPs found the point system easy to understand (Table 5).

### 3.3 Reflections on diabetes-specific experiences

#### 3.3.1 Emotional distancing through linking diabetes challenges to personas enabled articulation of reflections

Linking diabetes-specific issues to fictitious personas promoted emotional distancing that enabled PWT2D to express their individual experiences with diabetes. Stepping in and out of the personas was a gateway for PWT2D to articulate their reflections, as explained by one HCP: “Well, very quickly, they forget Jens and Jytte [names of fictitious personas], and then they talk about themselves and their own lives with diabetes.” (HCP, municipality I)

Eighty-six percent (62) of PWT2D and 81% (13) of HCPs agreed or strongly agreed that the fictitious personas seemed authentic, and 76% (58) of the PWT2D reported that the personas made them think of their own lives with T2D (Table 5).

#### 3.3.2 Confusing game rules led to uncertainty about how to play

Although playing personas helped some PWT2D articulate their reflections, other participants found the game structure unclear in terms of when to play as oneself and when to play as the persona. Thus, the groups occasionally spent time understanding the game rules instead of reflecting and participating in peer dialogues:

> P2: “I have to admit that I get a bit confused. Because, when is it exactly that I have to talk about Jens [the persona], and when do I have to talk about myself? Then, it becomes kind of a mess and I end up simply saying what I think.” (focus group 5)

### 3.4 Structured and focused dialogues

#### 3.4.1 Playing a fictitious persona structured an open dialogue

PWT2D mirror parts of themselves in the persona, as explained by one HCP: “It becomes kind of harmless when it’s based on a fictitious persona. Then, it really isn’t me, but her [the persona] that it’s about” (HCP, municipality E). Reading aloud the short text describing the fictitious persona and then explaining similarities and differences between the persona and the player’s own life situation was a structured and focused way to start an open dialogue.

Ninety-six percent (71), 88% (67) and 83% (62) of PWT2D agreed or strongly agreed that they had fruitful conversations, talked with others about diabetes and had listened to others’ experiences with diabetes, respectively. In addition, 83% (63) agreed or strongly agreed that they had experienced a sense of collectiveness with co-players, while 76% (N13) of the HCPs agreed or strongly agreed that the game encouraged dialogue within the group (Table 5).

#### 3.4.2 Choosing between different diabetes-specific topics supported PWT2D in sharing experiences

The theme cards were broadly related to everyday life and included emotional and social aspects of diabetes. The title, humorous picture and quote on each theme card was intended to summarise the essence of each theme, helping PWT2D quickly grasp the topics of theme cards and easily choose the best topic for themselves or their persona. This provided players with opportunities to share and address their experiences on meaningful and taboo topics, as expressed by one HCP:

> “The issue about guilt and shame [prompt on the theme card]. Two of the women were pretty affected by that, right? I knew that one of them was vulnerable, but not that much. Much of the stuff came up when playing the game. And when it comes up like that, then it means a lot to them. It’s good to know. You get to know them in a different way.” (HCP, municipality I)

Survey data revealed that 86% (65) of PWT2D agreed or strongly agreed the game topics were relevant to everyday life with T2D. On a visual analogue scale (VAS) of 0-10, 88% (15) HCPs rated the game ≥ 5 as promoting useful knowledge about individual diabetes-related experiences of PWT2D (Table 5).

#### 3.4.3 Lack of time rushed peer dialogue

Facilitating in-depth dialogue within the group was perceived as a challenge within the timeframe of the game, as expressed by one HCP: “It’s just a shame if you get stressed due to time [constraints], because it’s [engaging in dialogue] really important to them, as you can tell [from the focus group interview].” (HCP, municipality B)

### 3.5. Healthcare professionals gained insights into preferences and needs of people with type 2 diabetes

#### 3.5.1 Limited educator talk enhanced time for dialogues among peers

A set of rules framing ways to play the game encouraged HCPs to listen more and talk less. This provided the HCPs with insights into preferences and needs of PWT2D, as stressed by one HCP:

> “During the game, they [PWT2D] have to do the talking, not me. I think that’s what the game is very good at [supporting], because it contributes to creating a space where it’s more about them [PWT2D] and I’m less important. I’m not watching over them as much as I usually do.” (HCP, municipality B)

Questionnaire data showed that 59% (10) of HCPs reported that the game prompted PWT2D to talk more than usual in DSME programmes, and 88% (15) of HCPs agreed or strongly agreed that they had listened to the experiences of PWT2D during the game (Table 5).

#### 3.5.2 HCP uncertainty about facilitation inhibited dialogue

HCPs frequently mentioned feeling uncertain about their abilities to facilitate peer dialogues during the game. This led to a variety of strategies to facilitate dialogues. In one setting, HCPs chose to override the game structure by becoming the primary person that PWT2D paid attention to. The HCPs did not make room for or encourage peer support, as opposed to including participants in the dialogue by finding commonalities across their experiences: “Perhaps, I was very controlling in my group, but they needed some guidance. I don’t see how they could’ve played it without me being there” (HCP, municipality F). A few HCPs explained that they overrode the game structure due to scepticism and concern that playing the game was too time-consuming and resulted in less time available for other programme activities. In contrast, other HCPs explained how they remained in the background, guiding the dialogue only when needed to ensure that it stayed on track.

### 3.6 Healthcare professionals experienced peer dialogue as important to incorporate in education

#### 3.6.1 Playing the game created solidarity and a sense of collectiveness

HCPs expressed varying views of the importance of prioritizing peer dialogue and active engagement. Some found the game-induced peer dialogues crucial for PWT2D, as expressed by one HCP: “[…] it [the game] brought them closer together. I already sensed that during the game. That it created a kind of solidarity. It was an eye opener how much they actually needed to tell each other things and to talk to equals” (HCP, municipality I). Survey data revealed that 92% (70) of PWT2D and 94% (16) of HCPs experienced the game as good, very good or excellent. On a VAS scale of 0-10, 88% (15) of HCPs rated as ≥ 5 their desire to use the analogue game in their future work, and 88% (14) rated their likelihood of recommending the game to a colleague as ≥ 5 (Table 5).

#### 3.6.2 Used too much time in understanding the game

Although the questionnaire responses indicated that most HCPs would consider implementing the game in future DSME, others were unsure whether they would do so due to its time requirements:

> “[…] if we’re really supposed to have time for dialogues and reflections [when playing the game], then we need double the time, and then I think we could just as well have the dialogue in another way spending much less time […] they spend too much energy understanding the game instead of reflecting on how to live their lives with diabetes.” (HCP, municipality H)

Sixty-three percent (10) of HCPs agreed or strongly agreed that playing the game left too little time for other programme activities, and 50% (8) reported that it was difficult to end dialogues during game play (Table 5).

## 4. DISCUSSION

Our study provides novel insights into outcomes of using a structured framework to incorporate peer support and person-centredness in group-based DSME. By exploring hypothesized mechanisms in different settings, we gained an understanding of how the game worked, which can inform implementation in similar settings and may also be transferable to peer support and person-centredness in general. The analysis revealed game factors that either enabled or inhibited peer dialogue and person-centredness. As a playful activity, the game promoted a relaxed atmosphere of trust that, in combination with game rules, promoted structured and focused dialogues, encouraging PWT2D to share diabetes-specific experiences. In contrast, lack of time and complicated game rules somewhat inhibited peer dialogue and person-centredness.

Playing the game facilitated active engagement among players. An important game feature was engaging PWT2D who might otherwise have found it challenging to become actively involved. Other studies emphasize the importance of involving PWT2D in DSME programmes and the ability of dialogue tools to promote active involvement (Torenholt, Varming, et al., 2015; Varming et al., 2018). Difficulty engaging in DSME among PWT2D may be related to low levels of health literacy. Previous studies indicate that it may be especially important that PWT2D with low levels of health literacy receive the benefits of involvement and engagement in DSME programmes (Saunders et al., 2016; Torenholt, Varming, et al., 2015). This is consistent with the findings of Hartman et al. (1994), in which individuals with limited literacy preferred practical hands-on activities in educational programmes. Learning through games may be of particular value to this group.

The game provided multiple ways for participants to address their needs and share their experiences, which provided HCPs with detailed insights into needs and preferences of PWT2D. However, the game alone cannot promote a person-centred approach in self-management education. Its successful use depends on the ability of HCPs facilitating the game to incorporate participants’ preferences and needs into programme content (Torenholt, Engelund, et al., 2015). Most HCPs emphasized the value of the game in facilitating dialogue, reflection and active engagement among PWT2D. However, contextual factors related to HCP preconceptions about and rationales for implementing person-centredness and peer dialogue and imposing time constraints are key aspects to address when implementing the game in the future.

It can be extremely challenging for HCPs to incorporate person-centredness as part of diabetes care because doing so calls for a cultural change in practice (Joseph-Williams et al., 2017). The game in our study served as a structured format with a set of rules that facilitated inclusive and focused dialogues, encouraging HCPs to listen more and talk less. However, HCPs who overrode the game structure and dominated interactions inhibited peer dialogue. This is consistent with other studies showing that the potential for incorporating person-centeredness in DSME largely depends on the communication skills of HCPs and that their fundamental mindset must be addressed before specific tools are employed (Jensen et al., 2016; Stenov et al., 2019). These findings emphasize the importance of detailed introduction to the rationale behind the game rationale before HCPs use it. A review by Fisher et al. (L. Fisher et al., 2017) identified two crucial steps HCPs must complete before using dialogue tools. They must first be supported in shifting their perspective from a traditional hierarchical approach to a collaborative and empathic approach and from a traditional educational approach of delivering information toward listening. The second and equally fundamental step is to support HCPs in applying empathic relationship-building strategies. Using available tools is the final step in enhancing self-management.

### 4.1 Limitations

The primary limitation was that a researcher involved in developing the game and HCPs who facilitated the game both attended focus groups, potentially biasing participants’ evaluations. However, the questionnaire allowed participants to maintain anonymity and was conducted before the focus group. It is also unknown whether observed outcomes result in long-term benefits for PWT2D. A study strength is the large volume of data and triangulation with questionnaires, focus groups and interviews, as well as audio recording and observations of game sessions. Triangulation also revealed inconsistencies in the data, such as discrepancies between the responses of PWT2D and HCPs. Including urban and rural settings was intended to increase the variation in sociodemographic characteristics of participating PWT2D, increasing the potential transferability of the game to PWT2D with various backgrounds and across settings.

## 5. CONCLUSION

The analogue game served as a playful and structured format that supported HCPs in facilitating person-centeredness and peer dialogue in practice. Similar structured and playful formats can serve as useful frameworks to enhance person-centeredness and peer support in DSME programmes. However, the mindsets and communication skills of HCPs are crucial to facilitating person-centeredness and peer support in DSME programmes, even when employing structured formats. Methods to support HCPs in focusing on person-centeredness are needed.

## Data Availability

The data are not publicly available due to privacy or ethical restrictions.

## Acknowledgements

We are sincerely thankful to the participating people with type 2 diabetes and healthcare professionals for their valuable contributions to the initial workshops and for letting us test the analogue game in the diabetes self-management education programmes, followed by interviews and focus groups. We acknowledge Copenhagen Game Lab for collaborating in the development of the analogue game. Finally, we wish to thank student assistant, Thit Hjortskov Jensen, for her efforts in the data collection and data transcription processes, as well as student assistant, Maria Friis Børsting, for helping with the initial coding of data.

---

**Appendix 1.** Overview of the analogue game development process

| Purpose | Activities | Participants |
| --- | --- | --- |
| <u>Phase 1: Ideation</u> | Kick-off workshop for brainstorming ideas | Researchers and game designers |
| Define the approach and learning objectives | Interviews with domain stakeholders and specialists to generate ideas | Diabetologist and diabetes nurse |
| Develop the core game structure | Game-design workshop to generate ideas | Game designers |
| Develop early concept prototypes | Interactive co-creation workshops focusing on themes and methods for the game guided by principles to promote person-centredness and peer support | Researchers and game designers |
|  | Developing and iterating on the game structure, narrative and content using patient quotes and personas based on existing educational material | Researchers and game designers |
| <u>Phase 2: Development</u> | Fine-tuning the game structure | Game designers |
| Iteratively prototype, test and redesign the game based on observations and user feedback | Exploring reactions to preliminary prototypes from stakeholders by testing and adjusting prototypes in eight DSME sessions | 1-2 HCPs and 3-12 PWT2D per test from eight different settings across three municipalities |
|  | Adjusting prototypes based on user feedback, (e.g., developing a new point system, adding more pictures and condensing the text) |  |
|  | One test with external game designers and two tests with researchers | Game designers and researchers |
| Finalize graphic design, enhance usability and initiate manual production | Final workshops with researchers and game developers | Researchers and game developers |
|  | Writing the facilitator manual | Researchers |
|  | Developing graphic design | Graphic designer |
|  | Testing the game with researchers and game developers | Researchers and diabetes psychologist |
|  | Making final adjustments to contents and graphic design | Researchers and game designers |

